# Personalized CZA-ATM dosing against an XDR *E. coli* in liver transplant patients; the application of the *in vitro* hollow fibre infection model (HFIM)

**DOI:** 10.1101/2024.04.08.24301402

**Authors:** Zahra Sadouki, Emmanuel Q. Wey, Sateesh Iype, David Nasralla, Jonathan Potts, Mike Spiro, Alan Williams, Timothy D. McHugh, Frank Kloprogge

**Affiliations:** Institute for Global Health, University College London, UK; Centre of Clinical Microbiology, University College London, UK; Department of Infection, Royal Free London NHS Trust, UK; Department of HPB and Liver Transplant Surgery Royal Free Hospital; Department of Hepatology, Sheila Sherlock Liver Unit, Royal Free London; Department of Surgical biotechnology, UCL; Department of Intensive care medicine Royal Free London; Department of Infection Sciences, Health Services Laboratories, London, UK

**Author notes:** **Corresponding author:** Dr Emmanuel Wey, Department of Infection, Consultant and Honorary Associate Professor in Infection, Royal Free London NHS Trust, Centre for Clinical Microbiology, Division of Infection & Immunity, University College London. Both authors contributed equally (Zahra Sadouki, Emmanuel Q. Wey). **Financial support statement:** This work was conducted as part of Z.S.’s PhD studentship that was partially funded by an educational grant from Shionogi B.V. and by University College London. FK has been recipient of a UKRI Medical Research Council Skills Development Fellowship MR/P014534/1, and a Sir Henry Dale Fellowship jointly funded by the Wellcome Trust and the Royal Society (grant number 220587/Z/20/Z). **Authors contribution statement:** All authors contributed to the inception and planning of the work. ZS ^⍰^ and EW^⍰^ wrote the manuscript and contributed equally, and all authors edited.

**Keywords:** XDR *Escherichia coli*, Ceftazidime-Avibactam, Aztreonam, personalised medicine, hollow fibre infection model

## Abstract

**Background & aims:** An extensively-drug resistant (XDR) NDM and OXA-48 producing *E. coli* contributing to repeat episodes of biliary sepsis was isolated from the blood stream of a 45-55 year-old male with a background of IgG4 related sclerosing cholangitis. The patient was awaiting orthotopic liver transplant (OLT). There is no standardized antibiotic prophylaxis regimen however in line with the Infectious Diseases Society of America (IDSA) guidance an antibiotic prophylactic regimen of Ceftazidime-Avibactam (CZA) 2.5g TDS with Aztreonam (ATM) 2g TDS IV was proposed.

**Methods:** To inform the individualised pharmacodynamic outcome likelihood prior to prophylaxis dosing the hollow fibre infection model (HFIM) was applied to simulate the *in vivo* antibiotic exposures of the CZA-ATM regimen. The HFIM was inoculated with ∼10 x 10^5^ bacterial CFU/mL of the XDR *E. coli* strain and CFUs/mL were measured for a total of 120 hours to determine the *in vitro* PK/PD killing dynamics.

**Results:** A 4-log reduction in CFU/mL in the first ten hours of the regimen exposure was observed however the killing dynamics were slow and six eight-hourly infusions were required to reduce bacterial cells to below the limit of quantification. Thus, the HFIM supported the use of the regimen for infection clearance however highlighted the need for several infusions. Standard local practise is to administer prophylaxis antibiotics at induction of OLT however the HFIM provided data to rationalise earlier dosing therefore the patient was dosed at 24 hours prior to their OLT induction. The patient was subsequently discharged 8 days after surgery.

**Conclusions:** The HFIM provides a dynamic culture solution for informing individualised medicine by testing antibiotic combinations and exposures against the bacterial isolates cultured from the patient’s infection.

## Introduction

Antibiotic prophylaxis in the perioperative period is the standard of care and routinely prescribed during surgical procedures including solid organ transplantation (SOT). Although evidence suggests an overall benefit in reducing postoperative infections the standard of care regimen and duration vary between transplant centres^1–3^. Here we present the application of the Hollow Fibre Infection Model (HFIM) to inform perioperative personalised antimicrobial dosing in a complex clinical setting. It is anticipated that antimicrobial combination therapy will require case-by-case based selection, accounting for individual risk factors and local patterns of resistance, given the rising incidence of multi drug resistant infections in solid organ transplant donors and recipients. Systematic reviews and meta-regression analysis has indicated that a there is statistically significant risk associated with gastrointestinal-tract colonisation of carbapenem resistant enterobacterales (CRE) and subsequent infection with CRE^4^. Informing personalised dosing requires *in vitro* testing beyond MIC reporting which for XDR and PDR infections only serves to highlight the limited options of antimicrobials^5^. In these contexts, HFIM provides an *in vitro* tool which can inform perioperative dosing. The semi permeable hollow fibres retain the bacteria in the extra capillary space (ECS) whilst allowing for flow of nutrients and drugs. Therefore, the bacteria isolated from the patient can be challenged against dynamic *in vivo* mimicking antimicrobial exposures to investigate bacterial PD^6^.

## Materials/Patients and methods

### Information on clinical findings and diagnostic assessment

An inpatient 45-55 year-old male at University College London Hospitals NHS Foundation Trust (UCLH), also under the care of Hepatology and Hepatobiliary surgery teams at Royal Free London NHS Foundation Trust (RFL), previously diagnosed with IgG related sclerosing cholangitis4 presented with recurrent biliary sepsis over a three-month period. Source control of the infections was not achieved by empirical antibiotic treatment due to the presence of three metallic stents in his biliary tree and an internalised percutaneous biliary drain. Blood culture samples and testing of the perihepatic collection fluid confirmed several MDR NDM, and OXA-48 positive gram-negative bacterial strains (Supplementary Table 3) with a combined resistance across nine major antibiotic classes. Further complicating the patient’s clinical management was the damage to the liver and loss of synthetic function therefore at the time the patient was placed on the NHS waiting list for orthotopic liver transplantation (OLT). The Infectious Diseases Society of America (IDSA) guidance on the treatment of antimicrobial-resistant gram-negative infections particularly MBL-producing CRE (e.g., NDM, IMP, VIM) suggests intravenous three times daily dosing of 2.5g Ceftazidime-Avibactam (CZA) in addition to 2g aztreonam (ATM)^7^. In addition, several clinical infectious disease publications report efficacy of CZA plus ATM in patients with BSIs ^8–10^. Further antibiotic susceptibility testing (AST) revealed CZA-ATM synergy where ATM activity was restored when in combination with AVI (Supplementary Table 1). Therefore, the HFIM was employed as a proof of concept to simulate the *in vivo* dynamic concentrations of the proposed treatment of a three-way antibiotic dosing regimen to be administered to a clinically complex patient.

### Laboratory consumables and experimental setup

The laboratory control strain (ATCC 25922^®^) and the clinical isolate (XDR *E. coli*) were prepared identically. Each were cultured at 37°C in ambient air on nutrient agar and cryopreserved in Microbank™ beads at -80°C. MICs were determined following CLSI guidelines for microbroth dilution^11^. A mid-log phase inoculum for each isolate was prepared for the HFIM by refreshing a fresh overnight culture incubated in a shaking incubator at 200 rpm and 37°C in cation adjusted Mueller Hinton broth (CAMHB; Sigma, UK). A 1:50 dilution from this culture was performed to inoculate the HFIM ECS, achieving a final bacterial load of 10^5^ CFU/mL in the ECS.

The R-Shiny web application promoted by The International Society of Anti-Infective Pharmacology (ISAP) and ESCMID PK/PD of anti-infectives study group (EPASG) was used to define experimental setup parameters^12^. The following C_max_ concentrations were mimicked; CAZ 80 μg/L, AVI 14 μg/L, ATM 120 μg/L. A T_½_ of 2.48 hours was simulated in the HFIM. Antibiotic powders were dissolved in solvents recommended by the manufacturers (CAZ; Sigma, AVI; MCE and ATM; TOKU-E).

CAMHB (Sigma; UK) was broth media used in the HFIM system. The hollow fibre cartridge was sourced from FibreCell systems (High flux PS, C2011). The FibreCell duet pump was set at 100 mL/min to allow the cartridge and central reservoir to reach equilibrium. A peristaltic pump (Minipuls evolution^®^, Gilson) was set at 0.699 mL/min to mimic T_½_ of 2.48. The syringe driver pumps (AL-1000, WPI) were programmed to automatically dose 2 mL infusions every 8 hours over 2 hours and 1 hour respectively for CZA and ATM, administration was concurrent and direct into the central reservoir. Dosing timepoints were at 0, 8, 16, 24, 32, 40, 48, 56, 64, 72, 80, 88, 96, 104 and 112.

The cartridge ECS was sampled on Mueller Hinton agar (MHA) (VWR; UK) to quantify CFUs as well as on colombia blood agar (VWR; UK) to confirm the absence of contamination. The experiment with the laboratory control strain (ATCC 25922^®^) and the clinical isolate (XDR *E. coli*) were both performed in biological duplicate and all bacterial CFU/mL measurements were performed in technical duplicates at timepoints 0, 2, 4, 6, 8, 10 to capture the initial killing dynamics then again, every 24 hours for five days to capture sustained killing or re-growth.

## Results

MICs demonstrated synergy between ATM-AVI against the XDR *E. coli* isolated from the patients’ blood stream. MICs were as follows CAZ; >80, AVI; >14, ATM; >120, CZA; >32/4, ATM-AVI; 2/4 and CZA-ATM; 2.5/0.44/3.8. Although CLSI and EUCAST are yet to establish breakpoints for the ATM-AVI combination, this MIC and synergy data seen fell within range of previous *in vitro* studies reporting MICs between ≤0.03/4 and 8/4 μg/mL, most typically ≤2/4 μg/mL for MBL-producing Enterobacterales^13,14^ .

The XDR *E. coli* challenged with CZA-ATM combination therapy infusions produced a steady rate of killing for the first 10 hours with a 4-log reduction in bacterial CFU/mL however six eight-hourly infusions were required to reduce bacterial cells to below the limit of quantification (Figure 1). Therefore, multiple infusions were required to achieve bacterial source control. The HFIM data demonstrated the proposed combination therapy was only effective at clearing the XDR *E. coli* after two days of administration of eight hourly infusions. Thus, this suggests longer extended infusions or earlier administration of prophylaxis would be necessary for this patient as towards the end of the infusion the simulated concentration of both ATM and CAZ drop below their respective MICs.

**Figure 1.**
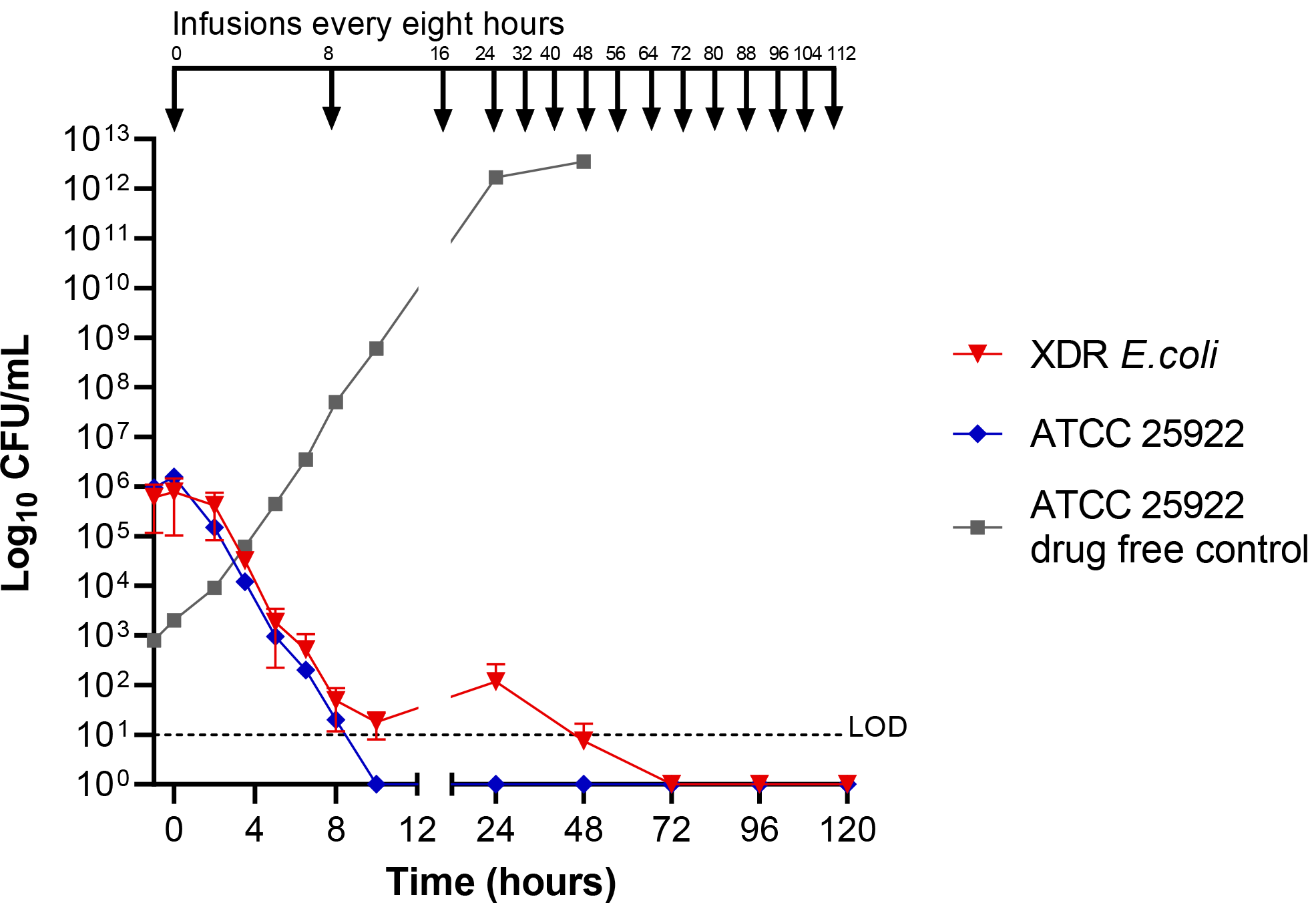
XDR *E. coli* against Ceftazidime/Avibactam/Aztreonam combination regimen in HFIM. Total bacterial population counts in CFU/mL over time. Data points for XDR *E. coli* (red) represent the geometric mean ±SD of biological duplicates. Antibiotic infusions were dosed every 8 hours (shown in black at the top of the graph). The lower LOD for quantifying the bacteria was 10 CFU/mL, shown as black dashed line.

In general clamping of the inferior vena cava occurs at two to three hours after induction followed by anhepatic period from three to four hours post induction of the OLT during which the mobilisation and explantation of the native Liver takes place. It would be at this point when and mobilisation and removal of the primary prosthetic sources of XDR biofilm would take place. Surgical implantation of the new deceased donor graft would therefore theoretically coincide with the MIC nadir and potential bacteraemia resulting from explantation of the native liver and if the first dose were given at induction it would also coincide a CFU/mL load above the LOD. This informed the patients antibiotic prophylaxis, and Ceftazidime avibactam 2.5g Iv and Aztreonam 2g IV were first dosed at 12 hours prior to the anhepatic phase of their OLT induction thus different to the standard local practise of administering prophylaxis antibiotics at induction of OLT 8-hoturly for a total period of 48 hours. The patient received a DCD graft with side-to-side piggyback anastomosis and portal vein end-to-end anastomosis and Roux-en-Y Hepaticojejunostomy with removal of a previously internalised percutaneous biliary drain.

The patient was discharged 8 days after surgery and had no reoccurrence of XDR *E. coli* infection reported in the year after follow up. Incidentally, the patient has also remained CRE negative on screening post OLT.

## Conclusions

Several attempts have been made to develop *in vitro* laboratory susceptibility testing methods for the combination of CZA with ATM against XDR GNB^15–17^. However, these methods test static antibiotic concentrations. Here we challenged a patient XDR BSI isolate against dynamic *in vivo* simulated concentrations of the CZA-ATM antibiotic combination. The HFIM data demonstrated the CZA-ATM antibiotic regimen was effective at clearing the bacterial cells below the limit of detection in the hollow fibre ECS however multiple infusions were required. The Royal Free London Trust standard of care is to administer prophylactic antibiotics for orthotopic liver transplant (OLT) at induction^3^. However, the slow killing dynamics observed in the HFIM provided *in vitro* evidence to support prophylactic administration prior to OLT as well as an extended prophylactic duration of 48-hours. Therefore, the patient awaiting a liver transplant was dosed 12 hours earlier than standard local practice. The patient was subsequently discharged 8 days after OLT. The unpredictability relating to the timing and lead times associated with deceased donor grafts offers complicates the practicality of commencing antibiotic prophylaxis in this patient cohort far in advance of 12 hours. This is compounded by the requirement to administer both ceftazidime-avibactam and aztreonam concomitantly.

The increased application of alternative synergistic combinations of antibiotic agents necessitates improved methodologies and approaches of *in vitro* evaluation of combination regimens. We demonstrate extended prophylaxis periods are superior for a chronically infected patient where source control has not been achieved with conventional antibiotic treatment and alternative combination regimens are used. The need of standardized methods to support personalised medicine will be increased as the antibiotic resistance era heightens. The HFIM provides a solution to testing different antibiotic regimens against the exact bacterial isolate cultured from the patient. Specific PK concentration profile of the proposed antibiotics can be mimicked. The increased prescribing of alternative synergistic combinations necessitates improved *in vitro* methodology for evaluating personalized dosing. The HFIM provides a dynamic culture solution for testing personalized antibiotic regimens and exposures against the bacterial isolates cultured from the patient. This could inform targeting therapy which could preserve antimicrobials and uphold antimicrobial stewardship.

## Data Availability

All data produced in the present study are available upon reasonable request to the authors

## Informed consent

The patient was consented for Orthotopic liver transplantation and normothermic perfusion as is standard for all patients and includes consent for Research publications of outcomes and the use of peri-transplant tissue samples for research. In this case no patient or deceased donor tissue was used in vitro testing.

## Notes

**Conflict of interest statement:** No conflicts of interest.

### Competing Interest Statement

The authors have declared no competing interest.

### Funding Statement

This work was conducted as part of Z.S. PhD studentship that was partially funded by an educational grant from Shionogi B.V. and by University College London. FK has been recipient of a UKRI Medical Research Council Skills Development Fellowship MR/P014534/1, and a Sir Henry Dale Fellowship jointly funded by the Wellcome Trust and the Royal Society (grant number 220587/Z/20/Z).

### Author Declarations

Health Research Authority Decision Panel advisors did not consider the work research but a deviation from standard of care that was purely for clinical needs. Review by an NHS Research Ethics Committee was therefore not required.

