## Supplemental data for "Personalized CZA-ATM dosing against an XDR *E. coli* in liver transplant patients; the application of the *in vitro* hollow fibre infection model (HFIM)"

### Mean inhibitory concentrations (MICs) observed

#### MICs observed against BSI isolate

| **Table S1. MICs observed for XDR *E. coli* isolate and NCTC**® **12241 type *E. coli* versus EUCAST clinical breakpoints and EUCAST quality control (QC) ranges.** MICs achieved for antibiotic/s tested are presented in columns one (XDR *E. coli*) and three (NCTC *E. coli*). MICs are shaded in red to represent resistant clinical breakpoints category ranges for Enterobacterales. All MICs for the reference laboratory *E. coli* strain (ATCC® 25922) fell within EUCAST quality control ranges. | | | | |
| --- | --- | --- | --- | --- |
| **Antibiotic/s** | ***E. coli* XDR isolate modal MIC**  **mg/L** | **EUCAST clinical breakpoint**  **mg/L** | ***E. coli* ATCC 25922® modal MIC mg/L** | **EUCAST ATCC 25922® QC ranges**  **mg/L** |
| **CAZ** | >80 | R (>4) | 0.16 | 0.06-0.5 |
| **AVI** | >14 | - | >14 | n/a |
| **ATM** | >120 | R (>4) | 0.125 | 0.06-0.25 |
| **CZA*** | >32/4 | R (>8/4) | 0.25/4 | 0.06-0.5 |
| **ATM-AVI*** | 2/4 | - | 0.125/4 | n/a |
| **CZA-ATM** | 2.5/0.44/3.8 | - | <0.31/0.05/0.47 | n/a |

*****Avibactam concentration fixed at 4 µg/mL. CAZ: ceftazidime, ATM: aztreonam, AVI: avibactam

#### MICs observed against all cultures isolated from the liver transplant patient

| Table S3. MICs μg/mL observed against cultures isolated from the liver transplant patient. Red represents isolates in the resistant category and green represents isolates in sensitive category as per EUCAST clinical breakpoints for Enterobacterales. Isolate ‘HFIM_1_BSI *E.* coli’ was used in the HFIM experiments presented in Figure 1. | | | | | |
| --- | --- | --- | --- | --- | --- |
| **Antibiotic** | **HFIM_1_BSI** | **HFIM_2_Drain** | **HFIM_3_Drain** | **HFIM_4_Drain** | **HFIM_5_Abscess** |
|  | ***E. coli*** | ***E. coli*** | ***E. coli*** | ***M. morganii*** | ***E. coli*** |
| Ampicillin | >8 | >8 | >8 | >8 | >8 |
| Amikacin | <=4 | <=4 | 8 | <=4 | 8 |
| Aztreonam | >16 | 8 | >16 | 4 | >16 |
| Co-Amoxiclav | >32/2 | >32/2 | >32/2 | >32/2 | >32/2 |
| Ceftazidime | >16 | >16 | >16 | 2 | >16 |
| Cefixime | >2 | >2 | >2 | >2 | >2 |
| Ciprofloxacin | >1 | >1 | >1 | >1 | >1 |
| Colistin | <=1 | <=1 | <=1 | >4 | <=1 |
| Cephalexin | >16 | >16 | >16 | >16 | >16 |
| Ceftriaxone | >4 | >4 | >4 | >4 | >4 |
| Cefuroxime | >8 | >8 | >8 | >8 | >8 |
| Ertapenem | >1 | <=0.25 | >1 | <=0.25 | <=0.25 |
| Cefepime | >16 | <=1 | >16 | 2 | >16 |
| Nitrofurantoin | <=16 | <=16 | <=16 | 128 | <=16 |
| Fosfomycin | <=16 | <=16 | <=16 | 64 | 32 |
| Gentamicin | >4 | 2 | >4 | >4 | - |
| Imipenem | >8 | <=0.25 | >8 | 8 | 2 |
| Levofloxacin | >2 | >2 | >2 | >2 | 1 |
| Mecillinam | >8 | <=2 | >8 | - | >2 |
| Meropenem | >8 | <=0.125 | >8 | 0.25 | 0.25 |
| Nalidixic acid | >16 | >16 | >16 | >16 | >16 |
| Tobramycin | >4 | 2 | >4 | >4 | >4 |
| Piperacillin | >8 | >64 | >8 | >8 | >8 |
| Norfloxacin | >64 | >2 | >64 | - | >64 |
| Co-trimoxazole | >4/76 | >4/76 | >4/76 | >4/76 | <=1/19 |
| Temocillin | >32 | 16 | >32 | - | >32 |
| Tigecycline | 1 | <=0.5 | 1 | 2 | 1 |
| Ticarcillin/clavulanate | >64/2 | >64/2 | >64/2 | >64/2 | >64/2 |
| Trimethoprim | >4 | >4 | >4 | >4 | >4 |
| Piperacillin/tazobactam | >64/4 | <=4/4 | >64/4 | <=4/4 | >64/4 |

### CZA-ATM concentrations simulated in the HFIM

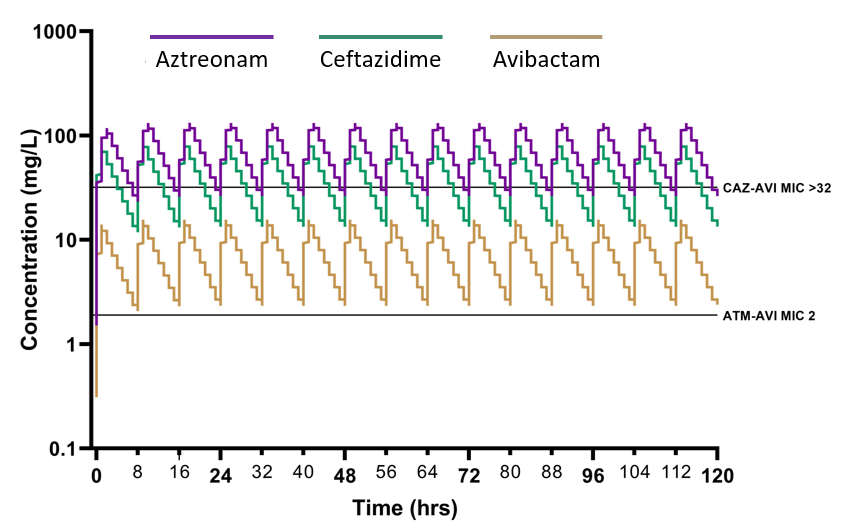

**Figure S1. HFIM simulated concentrations of CZA-ATM**

Dynamic concentrations of aztreonam (ATM), ceftazidime (CAZ) and avibactam (AVI) simulated in the HFIM experiments using *in vivo* PK data. Solid lines on the y axis represent the MIC for each antibiotic/combination tested in static microbroth dilution.

### HFIM methodology

| **Table S2. HFIM experimental set up including technical and microbiological specifications** | | |
| --- | --- | --- |
| Section | HFIM feature | Further explanation |
| Descriptive specifications | Primary aim | Mimic antibiotic combination therapy proposed for patient |
|  | Microbial species | XDR *E. coli* inoculated into extra-capillary space (ECS) |
|  | Antimicrobial/s | Ceftazidime + avibactam + aztreonam |
|  | Duration | 120 hours (5 days) |
| Technical specifications | Mimicked dose | 2.5g CZA over 2hr + 2g ATM over 1hr |
|  | System volume | 150 mL (central 80 mL + cartridge 70 mL) |
|  | C_max_ | Ceftazidime 80 µg/L (∴ administered 7.8 µg/L)  Avibactam 14 µg/L (∴ administered 1.4 µg/L)  Aztreonam 120 µg/L (∴ administered 1.4 µg/L) |
|  | T_1/2_ | 2.48 hours |
|  | Τ | 8 hours dosing interval |
|  | Drug administration | Syringe infusion (automated driver pump) |
|  | Cartridge source | FibreCell Systems Cartridge C2011 |
|  | Fibre type | High flux polysulfone |
|  | Pump model | FiberCell Systems® Inc duet pump at ~100 mL/min  Gilson pump set at 0.699 mL/min (5.92 rpm) |
| Microbiological specifications | Media | Mueller Hinton Broth 2, Cation-Adjusted (Sigma; 90922) |
|  | Control | Drug free experiment conducted in duplicate |
|  | Contamination | Sampling of ECS and reservoirs on CBA (VWR; 89407-240) |
|  | Inoculum | Initial OD600 for approximation then CFU/mL quantified |
|  | CFUs | CFUs quantified on MHA (VWR; 100611ZA) |
|  | Biological repeat | Duplicates |
|  | Technical repeat | Duplicates |
